# Accounting for imported cases in estimating the time-varying reproductive number of COVID-19

**DOI:** 10.1101/2021.02.09.21251416

**Authors:** Tim K. Tsang, Peng Wu, Eric H. Y. Lau, Benjamin J. Cowling

## Abstract

**Background:** Estimating the time-varying reproductive number, *R*_*t*_, is critical for monitoring transmissibility of an emerging infectious disease during outbreaks. When local transmission is effectively suppressed, imported cases could substantially impact transmission dynamics.

**Methods:** We developed methodology to estimate separately the *R*_*t*_ for local cases and imported cases, since certain public health measures aim only to reduce onwards transmission from imported cases. We applied the framework to data on COVID-19 outbreaks in Hong Kong.

**Results:** We estimated that the *R*_*t*_ for local cases decreased from above one in the early phase of outbreak to below one after tightening of public health measures. Assuming the same infectiousness of local and imported cases underestimated *R*_*t*_ for local cases due to control measures targeting travelers.

**Conclusions:** When a considerable proportion of all cases are imported, the impact of imported cases in estimating *R*_*t*_ is critical. The methodology described here can allow for differential infectiousness of local imported cases.

## INTRODUCTION

During an emerging infectious disease epidemics, monitoring transmissibility is important for guiding the implementation of public health measures and providing situational awareness on the effectiveness of interventions [1-3]. This can be achieved by estimating the time-varying reproductive number, *R*_*t*_, a measure of the transmissibility of the virus over time. A wide range of methods have been proposed [3-8] but the impact of imported cases on estimation of *R*_*t*_ is rarely explored [5].

Imported cases are cases that have been infected outside an area but travelled into and identified in the local area during their incubation period or symptomatic period. They have not been infected locally, but could potentially generate local transmissions. It is expected that the impact of imported cases on estimation of *R*_*t*_ would be limited when a local epidemic is underway as most infections occur locally. However, imported cases can play a more important role in the early stages of a local epidemic, or when local transmission is being suppressed by public health measures. While one study [5] proposed a modification by assuming the same infectiousness among imported and local cases, such modification may not be accurate if there are specific interventions targeting imported cases, such as testing on arrival at the border, or quarantine for inbound travelers [9].

To address such issues related to imported cases, we extend the framework in Cori et al [4] to estimate the *R*_*t*_ for local cases and imported cases separately. Using the first two waves of COVID-19 outbreaks in Hong Kong as an example, we illustrate the use of inappropriate methods to estimate *R*_*t*_ would lead to a biased assessment of transmissibility.

## METHODS

### Sources of Data

COVID-19 has been a notifiable disease in Hong Kong since 8 January 2020. Data on laboratory-confirmed COVID-19 cases were obtained from the webpage of Centre for Health Protection. Cases are classified as “imported cases”, “local cases epidemiologically linked with imported cases”, “unlinked local cases” and local cases epidemiologically linked with local cases” according to their epidemiological characteristics and location of infection [10].

### Statistical analysis

We examined different methods to handle imported cases in the estimation of *R*_*t*._ First, when we ignored the fact that imported cases were infected overseas and assumed them as local cases, *R*_*t*._ could be estimated in the framework described by Cori et al. [4]. In this approach, *R*_*t*_ was the ratio between the number of new cases at time *t* and the total infectiousness of cases at time *t*, given by 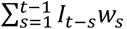 where *I*_*t*_ was the number of infections at time *t* and *w*_*t*_ was probability distribution of infectiousness profile. Second, assuming equal infectiousness for imported cases and local cases [5], the total infectiousness of cases could be represented by 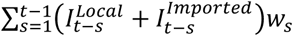, where 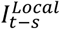 and 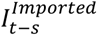 were the numbers of local and imported cases at time *t-s*.

To account for the different infectiousness of local cases and imported cases due to travel-related measures [11], we extended the framework to estimate the *R*_*t*_ of local cases and imported cases. We modified the estimation so that *R*_*t*_ for imported cases was the ratio of the number of new local cases infected by imported cases at time *t* and the total infectiousness of imported case at time *t*, given by 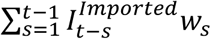. The *R*_*t*_ for local cases could be calculated by using the same approach.

For unlinked local cases, their source of infection was unclear. In addition, because of the potential for pre-symptomatic infectiousness of COVID-19 [12], it is preferable to conduct inference on the epidemic curve by infection date (unobserved) instead of by case confirmation date (observed). Therefore, we used a bootstrap approach that involved sampling of the source of unlinked local cases (linked with local cases or imported cases), and a deconvolution approach [13] to reconstruct the epidemic curve by infection date from the epidemic curve by confirmation date, with the distribution of delay from infection to confirmation, which was the convolution distribution of the incubation period distribution, with mean 5.2 days (SD 3.9) [14], and the empirical distribution of delay from onset to reporting. Our inference was based on a Bayesian framework and we developed Markov chain Monte Carlo algorithm to estimate the model parameters (Appendix). We conducted simulation to validate that our approach could obtain unbiased estimates of *R*_*t*_ for imported and local cases.

We used these three approaches to fit the data for the first two waves of COVID-19 outbreak in Hong Kong. We compared the estimates of *R*_*t*_ from the alternative methods. All analyses were conducted in R version 3.5.2 (R Foundation for Statistical Computing, Vienna, Austria).

## RESULTS

From 18 January to 8 May 2020, there were 719 imported cases and 326 local cases detected in Hong Kong. Among the 326 local cases, there were 62 (19%), 73 (22%) and 191 (59%) cases epidemiologically linked with imported cases, unlinked local cases, and epidemiologically linked with local cases respectively (Figure 1A).

**Figure 1.**
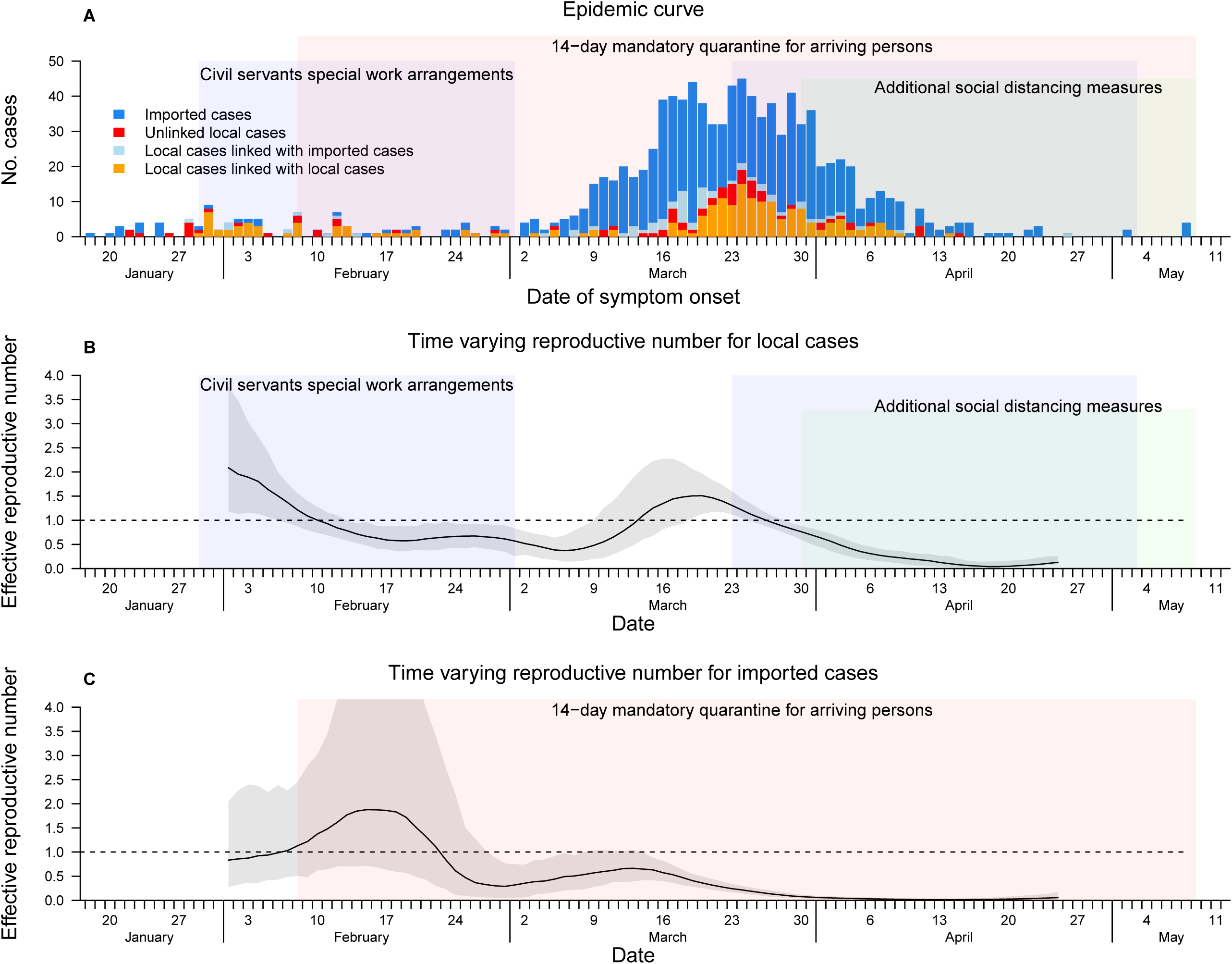
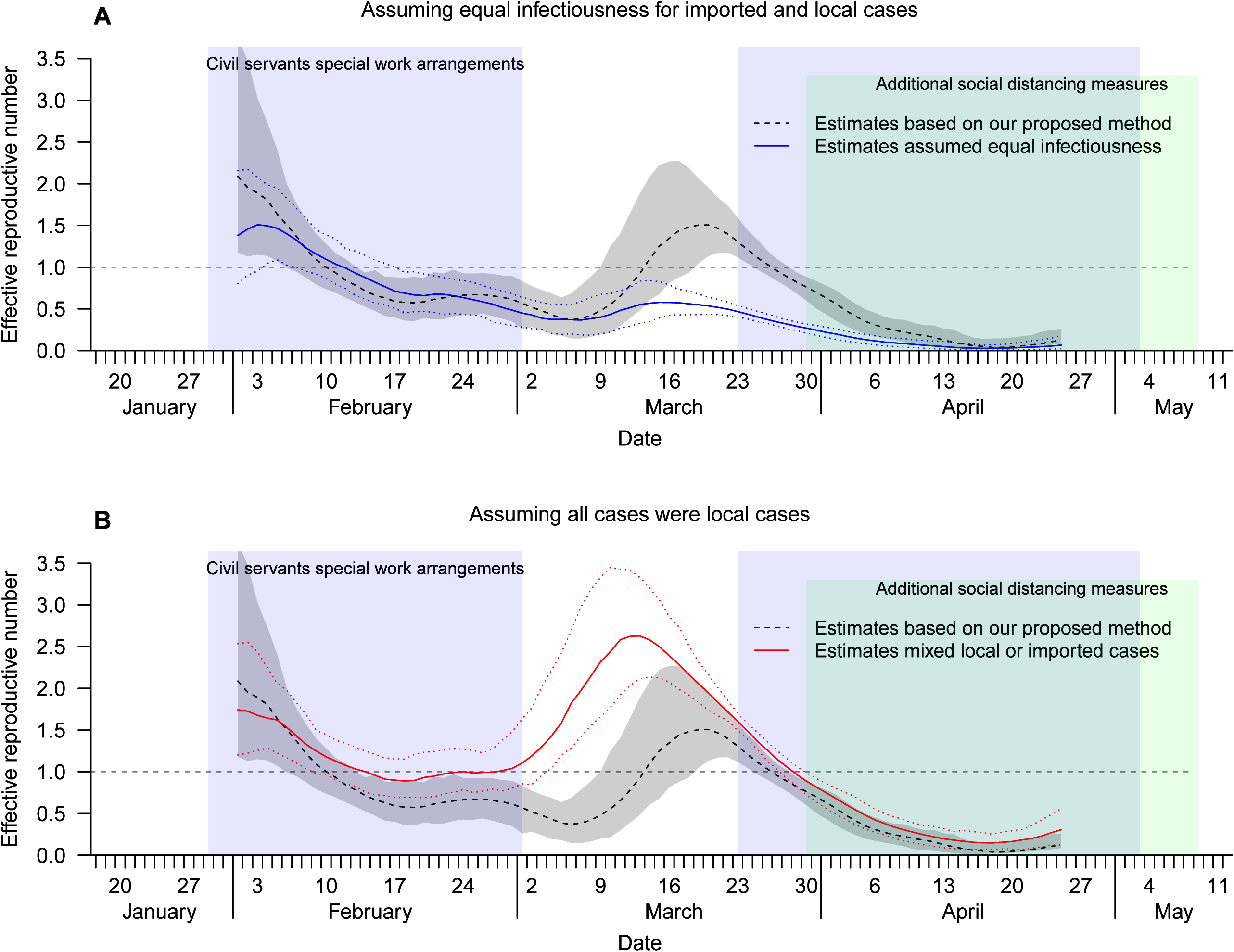
Epidemic curve of the COVID-19 transmission in Hong Kong from January through May 2020 (Panel A) and the estimated time-varying reproductive number for local cases (Panel B) and imported cases (Panel C).

Based on our approach, we estimated that the surge in local case numbers corresponded to an estimated *R*_*t*_ for local cases greater than one during March 14 to 26, which was the period before public health measures were tightened (Figure 1B). During the period with implemented interventions in the community (civil servant special work arrangement and additional social distancing measure), the estimated *R*_*t*_ for local cases was decreasing. The *R*_*t*_ for imported cases were well below one even when there were more than 10 daily imported cases during early March (Figure 1C), consistent with the implementation of a 14-day quarantine for inbound travelers.

We explored two alternative approaches to account for imported cases when estimating *R*_*t*_. Assuming same infectiousness for local cases and imported cases would have *R*_*t*_ less than one all along since late February, which would miss the local outbreaks in mid-March (Figure 2A). Assuming all cases were local cases in the estimation led to a peak in *R*_*t*_ in early March, which was inconsistent with the low number of local cases in that period (Figure 2B).

**Figure 2.**
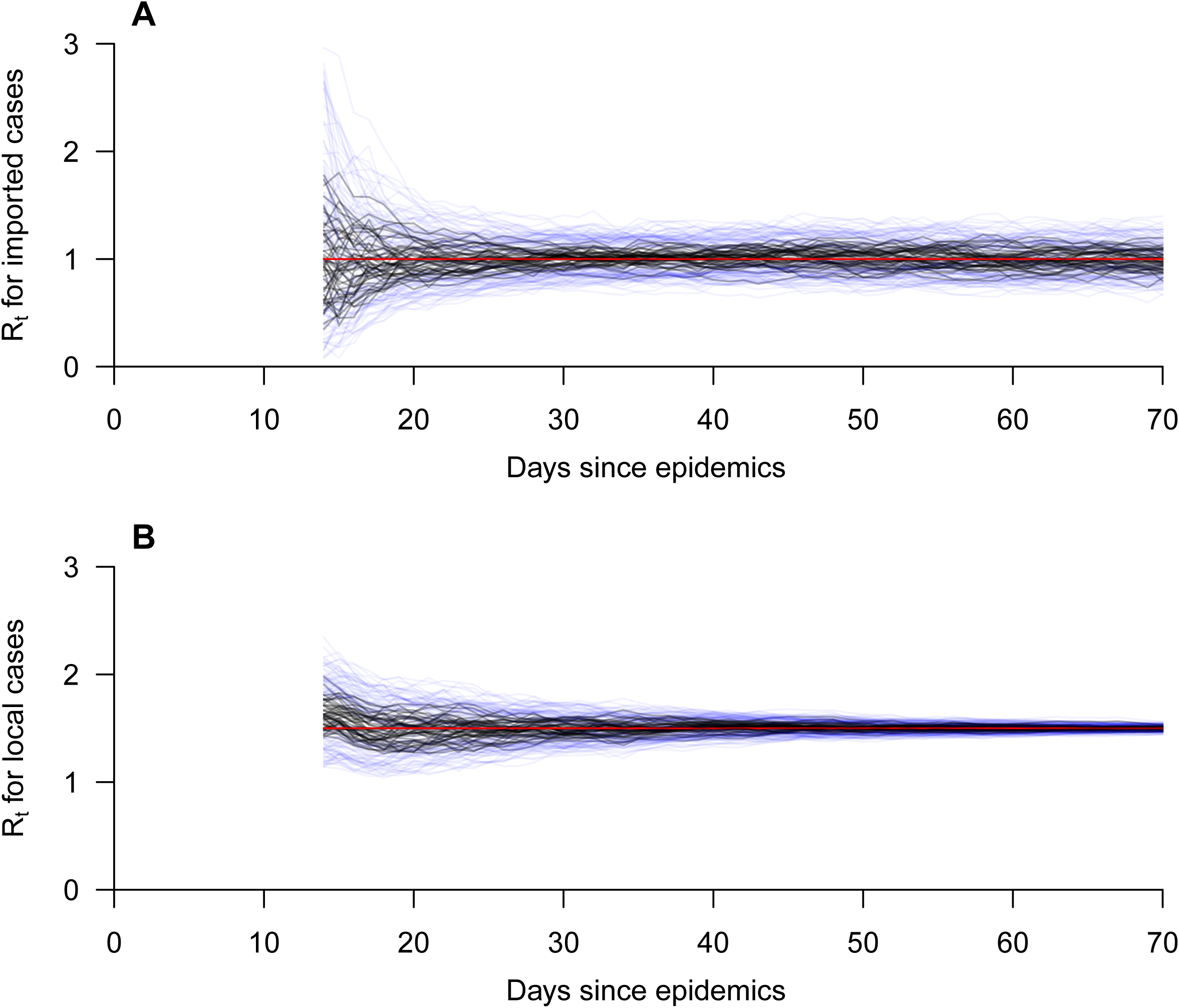
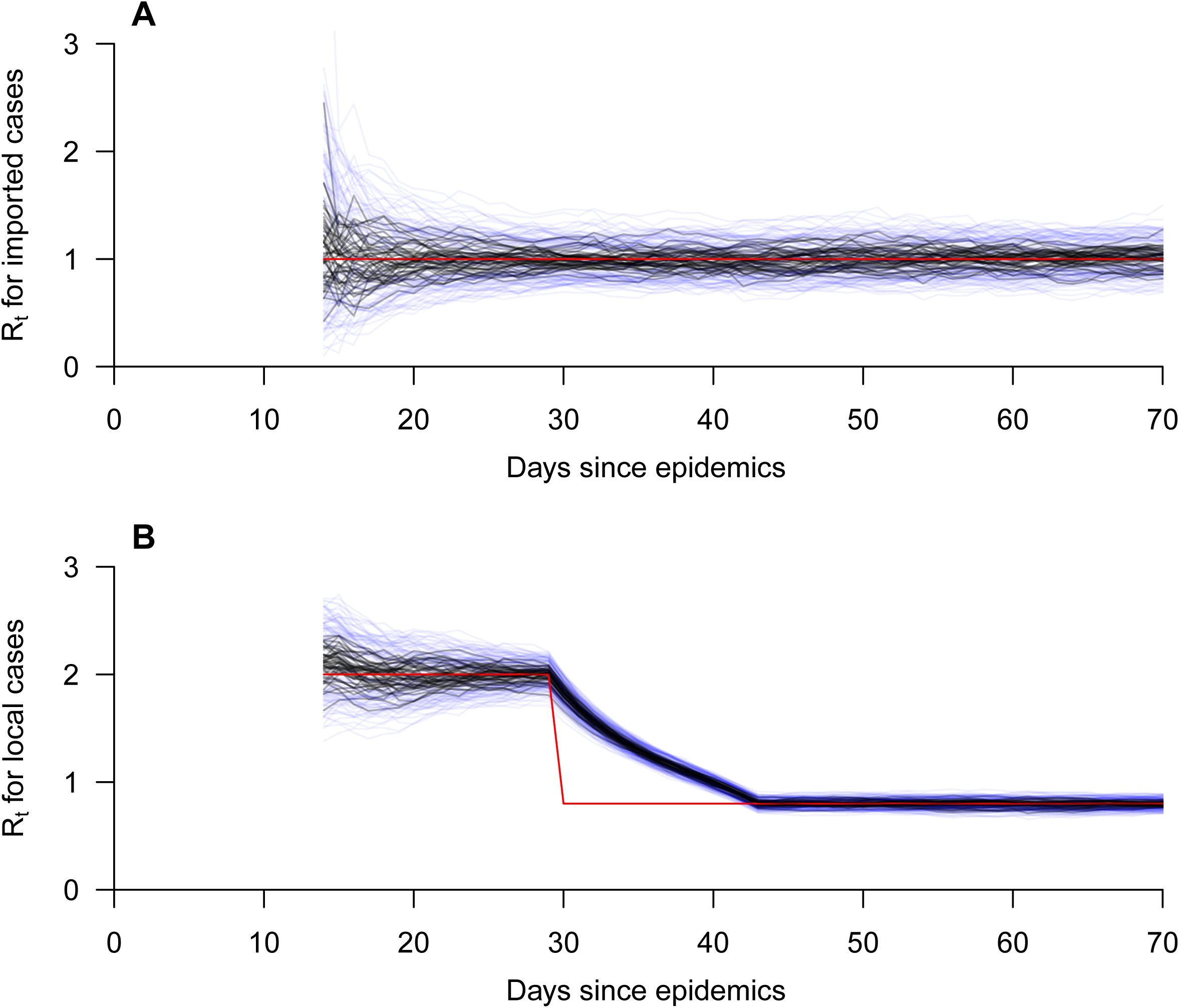
Comparison of estimation of time-varying reproductive number using our proposed framework, with alternative approaches that assuming equal infectiousness for local cases and imported cases (Panel A), or assuming all cases were local cases in the analysis (Panel B).

Simulation studies suggested that our model could obtain unbiased estimate of *R*_*t*_ for local cases and imported cases with constant *R*_*t*_ (Figure S1), and with an intervention controlling for local cases only so that *R*_*t*_ decreased from 2.0 to 0.8 (Figure S2).

## DISCUSSION

In this study, we extended the current framework for estimation of *R*_*t*_ to account for differential transmission from imported cases. In this framework, we separately estimated the *R*_*t*_ for local cases and imported cases, so that when control measures were targeting the imported cases, our framework would be feasible to allow *R*_*t*_ for imported cases to be lower than for local cases.

We compared the extended framework with the current two approaches, namely assuming all cases were local cases, or assuming equal infectiousness for imported cases and local cases [5]. These two approaches would have little impact when local transmission was substantial, but would bias the estimated *R*_*t*_ in measuring transmissibility in the community, when the number of local cases and imported cases were comparable. In particular, assuming all cases were local cases would overestimate the number of local transmissions as imported cases could not be infected locally. This was improved in the framework assuming equal infectiousness of local and imported cases [5]. However, this would be inappropriate if there were targeted control measures for inbound travelers.

Given that pre-symptomatic transmission was substantial for COVID-19, using serial intervals as a proxy of infectiousness profile would be inappropriate [4, 15]. Hence, we used a deconvolution approach to infer the epidemic curve by infection date from the observed epidemic curve that was by confirmation date, and using the infectiousness profile since infection [16]. Misspecification of the infectiousness profile since infection, or using a wrong approach to obtain the epidemic curve by infection dates such as back-shifting based on the delay distribution would bias the estimated *R*_*t*_ [8, 17]. We also properly accounted for the uncertainty in each step by using a bootstrap approach [18].

Our study has some limitations. First, we did not account for incomplete observations. Mild or asymptomatic cases would likely be undetected. Also, testing availability and criteria would also be likely changing over time. If the proportion of undetected cases were constant over the outbreak, the *R*_*t*_ would still be unbiased [17]. Further investigation would be necessary to develop method to account for the changing proportion of undetected cases. Finally, rapid changes in the parameters could affect the estimated *R*_*t*_, such as the sudden change for the delay distribution from infection to confirmation caused by overwhelmed healthcare systems, changing case definition during the epidemic [19], or superspreading events during the outbreaks [20].

In conclusion, we developed methodology to estimate separately the *R*_*t*_ for local and imported cases, to account for the potential differential infectiousness between them, due to control measures targeting inbound travelers. We compared the developed framework with the two current approaches to account for imported cases and illustrate the importance of properly accounting for the differential infectiousness in the estimation of *R*_*t*_. Accurate estimation of *R*_*t*_ allows situational awareness of transmission and control in the community.

## Data Availability

Data on the incidence of coronavirus disease 2019 and influenza activity are freely available from the Centre for Health Protection website (https://www.chp.gov.hk/en/index.html). Computing code could be obtained by contacting the corresponding author.

## ACKNOWLEDGMENTS

The authors thank Stephanie Gao and Faith Ho for technical assistance.

## REFERENCES

1. Ferguson NM, Cummings DA, Fraser C, Cajka JC, Cooley PC, Burke DS. Strategies for mitigating an influenza pandemic. Nature. 2006;442(7101):448–52. doi: 10.1038/nature04795. PubMed PMID: 16642006.

2. Fraser C, Riley S, Anderson RM, Ferguson NM. Factors that make an infectious disease outbreak controllable. Proc Natl Acad Sci U S A. 2004;101(16):6146–51. Epub 2004/04/09. doi: 10.1073/pnas.0307506101. PubMed PMID: 15071187; PubMed Central PMCID: PMCPMC395937.

3. Cauchemez S, Boelle PY, Thomas G, Valleron AJ. Estimating in real time the efficacy of measures to control emerging communicable diseases. Am J Epidemiol. 2006;164(6):591–7. Epub 2006/08/05. doi: 10.1093/aje/kwj274. PubMed PMID: 16887892.

4. Cori A, Ferguson NM, Fraser C, Cauchemez S. A new framework and software to estimate time-varying reproduction numbers during epidemics. Am J Epidemiol. 2013;178(9):1505–12. doi: 10.1093/aje/kwt133. PubMed PMID: 24043437; PubMed Central PMCID: PMCPMC3816335.

5. Thompson RN, Stockwin JE, van Gaalen RD, Polonsky JA, Kamvar ZN, Demarsh PA, et al. Improved inference of time-varying reproduction numbers during infectious disease outbreaks. Epidemics. 2019;29:100356. Epub 2019/10/19. doi: 10.1016/j.epidem.2019.100356. PubMed PMID: 31624039; PubMed Central PMCID: PMCPMC7105007.

6. Wallinga J, Teunis P. Different epidemic curves for severe acute respiratory syndrome reveal similar impacts of control measures. Am J Epidemiol. 2004;160(6):509–16. Epub 2004/09/09. doi: 10.1093/aje/kwh255. PubMed PMID: 15353409; PubMed Central PMCID: PMCPMC7110200.

7. Bettencourt LM, Ribeiro RM. Real time bayesian estimation of the epidemic potential of emerging infectious diseases. PLoS One. 2008;3(5):e2185. Epub 2008/05/15. doi: 10.1371/journal.pone.0002185. PubMed PMID: 18478118; PubMed Central PMCID: PMCPMC2366072.

8. Abbott S HJ, Thompson RN et al. Estimating the time-varying reproduction number of SARS-CoV-2 using national and subnational case counts [version 2; peer review: awaiting peer review]. Wellcome Open Res 2020, 5:112 (https://doi.org/10.12688/wellcomeopenres.16006.2).

9. Wells CR, Sah P, Moghadas SM, Pandey A, Shoukat A, Wang Y, et al. Impact of international travel and border control measures on the global spread of the novel 2019 coronavirus outbreak. Proc Natl Acad Sci U S A. 2020;117(13):7504–9. Epub 2020/03/15. doi: 10.1073/pnas.2002616117. PubMed PMID: 32170017; PubMed Central PMCID: PMCPMC7132249.

10. Cowling BJ, Ali ST, Ng TWY, Tsang TK, Li JCM, Fong MW, et al. Impact assessment of non-pharmaceutical interventions against coronavirus disease 2019 and influenza in Hong Kong: an observational study. Lancet Public Health. 2020;5(5):e279–e88. Epub 2020/04/21. doi: 10.1016/S2468-2667(20)30090-6. PubMed PMID: 32311320; PubMed Central PMCID: PMCPMC7164922.

11. Fraser C. Estimating individual and household reproduction numbers in an emerging epidemic. PLoS One. 2007;2(8):e758. doi: 10.1371/journal.pone.0000758. PubMed PMID: 17712406; PubMed Central PMCID: PMCPMC1950082.

12. Nishiura H, Linton NM, Akhmetzhanov AR. Serial interval of novel coronavirus (COVID-19) infections. Int J Infect Dis. 2020;93:284–6. Epub 2020/03/08. doi: 10.1016/j.ijid.2020.02.060. PubMed PMID: 32145466; PubMed Central PMCID: PMCPMC7128842.

13. Becker NG, Watson LF, Carlin JB. A method of non-parametric back-projection and its application to AIDS data. Stat Med. 1991;10(10):1527–42. Epub 1991/10/01. doi: 10.1002/sim.4780101005. PubMed PMID: 1947509.

14. Li Q, Guan X, Wu P, Wang X, Zhou L, Tong Y, et al. Early Transmission Dynamics in Wuhan, China, of Novel Coronavirus-Infected Pneumonia. N Engl J Med. 2020. doi: 10.1056/NEJMoa2001316. PubMed PMID: 31995857.

15. Wallinga J, Lipsitch M. How generation intervals shape the relationship between growth rates and reproductive numbers. Proc Biol Sci. 2007;274(1609):599–604. Epub 2007/05/04. doi: 10.1098/rspb.2006.3754. PubMed PMID: 17476782; PubMed Central PMCID: PMCPMC1766383.

16. He X, Lau EHY, Wu P, Deng X, Wang J, Hao X, et al. Temporal dynamics in viral shedding and transmissibility of COVID-19. Nat Med. 2020;26(5):672–5. doi: 10.1038/s41591-020-0869-5. PubMed PMID: 32296168.

17. Gostic KM, McGough L, Baskerville EB, Abbott S, Joshi K, Tedijanto C, et al. Practical considerations for measuring the effective reproductive number, Rt. PLoS Comput Biol. 2020;16(12):e1008409. Epub 2020/12/11. doi: 10.1371/journal.pcbi.1008409. PubMed PMID: 33301457; PubMed Central PMCID: PMCPMC7728287.

18. Salje H, Cummings DAT, Rodriguez-Barraquer I, Katzelnick LC, Lessler J, Klungthong C, et al. Reconstruction of antibody dynamics and infection histories to evaluate dengue risk. Nature. 2018;557(7707):719–23. doi: 10.1038/s41586-018-0157-4. PubMed PMID: 29795354; PubMed Central PMCID: PMCPMC6064976.

19. Tsang TK, Wu P, Lin Y, Lau EHY, Leung GM, Cowling BJ. Effect of changing case definitions for COVID-19 on the epidemic curve and transmission parameters in mainland China: a modelling study. Lancet Public Health. 2020;5(5):e289–e96. Epub 2020/04/25. doi: 10.1016/S2468-2667(20)30089-X. PubMed PMID: 32330458; PubMed Central PMCID: PMCPMC7173814.

20. Adam DC, Wu P, Wong JY, Lau EHY, Tsang TK, Cauchemez S, et al. Clustering and superspreading potential of SARS-CoV-2 infections in Hong Kong. Nat Med. 2020;26(11):1714–9. Epub 2020/09/19. doi: 10.1038/s41591-020-1092-0. PubMed PMID: 32943787.

