## Appendix for "Accounting for imported cases in estimating the time-varying reproductive number of COVID-19"

### 1.1 Summary of previous works

The time-varying reproductive number is discussed in Fraser [1]. There are two types of time-varying reproductive number, $R_t$, namely instantaneous $R_t$ and case $R_t$. It is showed that instantaneous $R_t$ could be straightforward to be estimated in real-time, because it does not involve assumption on future transmissibility. Therefore, we focus on using instantaneous $R_t$ to measure time-varying transmissibility.

Cori et al [2] is one of the first studies to estimate the $R_t$ from real data. In brief, it assumes that the distribution of infectiousness through time after infection is independent of calendar time. Transmission then is modeled by using a Poisson process. If $w_s$ is a probability distribution (sum equal to 1) of the infectiousness profile after infection, then the rate for infection at time step *t-s* generates new infections in time step $t$ is equal to $R_t w_s$, where $R_t$ is the instantaneous reproductive number at $t$. Also, the incidence at time $t$ is Poisson distributed with mean $R_t \sum_{s=1}^{t} I_{t-s} w_s$. It should be noted that we could use serial interval for proxy of infectiousness after symptom onset for inference [2], for disease with no or limited pre-symptomatic transmission.

Thompson et al. [3] then extends the method to account for imported cases by adjusting the mean by $R_t \sum_{s=1}^{t} \{I_{t-s}^{Imported} + I_{t-s}^{local}\} w_s$. However, such estimate assumed the average transmissibility of imported cases and local cases are the same and they are well-mixed in the population, which is unlikely to be true. In particular, this assumption is violated if there are interventions such as screening of travellers and quarantine measure.

To summarize, the following issues need to be addressed on estimating $R_t$:

1) Infection time is unobserved and hence estimation is based on symptom onset time. Particularly this would be an issue for disease with pre-symptomatic transmission.

2) Imported cases were not mixed well with the local population and their transmissibility may also be different, since they may be tested, quarantined or isolated due to travel-related measures. Therefore, they may have higher probability to be detected.

### 1.2 Model

Denote $Y_j(k)$ the number of infection at day $k$ for type $j$ (1: imported, 2: local case linked with imported cases, 3: local case linked with other local cases). Then, we have:

$$Y_2(t) \sim Poisson\{R_I(t) \sum_{k=1}^{t-1} Y_1(k) w_I(t-k)\}$$

$$Y_3(t) \sim Poisson\{R_L(t) \sum_{k=1}^{t-1} \{Y_2(k) + Y_3(k)\} w_L(t-k)\}$$

where $R_I(t)$ and $R_L(t)$ the time-varying effective reproductive number of imported cases and local cases at time $t$ respectively.

### 1.3 Likelihood function

We used the smoothing method as in Cori et al., meaning that the transmissibility is assumed constant over a time period $[t-\tau+1, t]$, where $\tau$ is the smoothing parameter. Hence likelihood at a time period $t$ is

$$P\big(Y_2(t), \dots, Y_2(t-\tau+1), Y_3(t), \dots, Y_3(t-\tau+1) \big| Y_2(1), \dots Y_2(t-\tau), Y_3(1), \dots Y_3(t-\tau)\big)$$

$$= \prod_{s=t-\tau+1}^{t} \frac{\big(R_L^\tau(t)\Phi_\mathrm{L}(s)\big)^{Y_3(s)} e^{-R_L^\tau(t)\Phi_\mathrm{L}(s)}}{Y_3(s)!} * \frac{\big(R_I^\tau(t)\Phi_\mathrm{I}(s)\big)^{Y_2(s)} e^{-R_I^\tau(t)\Phi_\mathrm{I}(s)}}{Y_2(s)!}$$

where $\Phi_\mathrm{L}(t) = \sum_{k=1}^{t-1}\{Y_2(k) + Y_3(k)\} w_L(t-k)$ and $\Phi_\mathrm{I}(t) = \sum_{k=1}^{t-1} Y_1(k) w_I(t-k)$. The total likelihood is the product of time period in the observed data, excluding for the first $\tau - 1$ days due to $\tau$-day smoothing.

### 1.4 Priors

For the parameters that only take positive values, such as standard derivation, we used a vague Uniform(0,10) prior. Otherwise, we used a vague Uniform(-

10,10) prior. We assumed the prior for $R(t)$ is Gamma(1,2) with mean and standard deviation equal to 2.

#### 1.5 Algorithm

At each MCMC step $k$, we update the model parameters $\theta$ by using random walk Metropolis-Hastings algorithm [4]. The step size of the proposal was adjusted to have acceptance rate for 20-30%.

#### 1.6 Simulation study

To validate that our approach could provide unbiased estimates of $R_t$ for imported cases and local cases. We conducted a simulation study. In each replication, we simulated an epidemic according to the input value of $R_t$ for imported cases and local cases, and the model in Section 1.2. Then we used our approach to estimate the unknown parameters.

In the first scenario, the $R_t$ for imported cases was set to be 1 and $R_t$ for local cases was set to be 1.5, with a 70-day epidemics. We summarized the estimates in Figure S1. In the 50 replications, most of the mean value (black lines) were close to the true value (red lines), and the 95% credible intervals (the blue lines) bracketed the true value, suggesting that our model provide a reasonable fit. As suggested in previous analyses [5, 6], the first 14 days estimates were ignored because they are biased.

In the second scenario, the $R_t$ for imported cases was set to be 1 and $R_t$ for local cases were set to be 2.0 in first 30 days, and then decreased to 0.8 for 40 days, mincing interventions that could control the local spread. We summarized the estimates in Figure S2. In the 50 replications, most of the mean value (black lines) were close to the true value (red lines), and the 95% credible intervals (the blue lines) bracketed the true value, suggesting that our model provide a reasonable fit. Although it shows that about 10 days were needed for the estimated $R_t$ to reach the value of post-intervention $R_t$.

### 2 Data analysis for Hong Kong COVID-19 outbreak

Since the time series of cases by infection dates could not be observed, we need to infer the epidemic curve by infection dates, to perform estimation. We also need to account for the uncertainty in each step by using a bootstrap type analysis.

### 2.1 Inference of epidemic curve by infection date

We used deconvolution approach in Becker et al. [7] to obtain the epidemic curve by infection time from the epidemic curve by onset time, and a given distribution of delay from infection to report. We summarize the approach as follows:

Given a known distribution of the delay from infection to report $f_{delay}(.)$, where

$$f_{delay}(u) = P(delay\ from\ infection\ to\ report = u)$$

Denote $Z_j(k)$ the number of onset at day $k$ for type $j$ (1: imported, 2: local case linked with imported cases, 3: local case linked with other local cases). Then, we have:

$$E\left(Z_j(k) \middle| Y_j(1), \dots, Y_j(k)\right) = \sum_{u=1}^{k-1} Y_j(u) * f_{delay}(k-u)$$

Denote $\mu_k = E(Z_j(k))$ and $\lambda_k = E(Y_j(k))$, we have

$$\mu_k = \sum_{u=1}^{k-1} \lambda_u * f_{delay}(k-u)$$

Under the assumption that $Y_j(k)$ are independent Poisson variates, $Z_j(k)$ are also independent Poisson variates. Hence, the likelihood function is

$$\prod_k \left[\sum_{u=1}^{k-1} \lambda_u * f_{delay}(k-u)\right]^{Z_j(k)} \exp\{-\sum_{u=1}^{k-1} \lambda_u * f_{delay}(k-u)\}$$

### 2.2 Assumption on input parameter in data analysis

For incubation period distribution, we use the estimate mean 5.2 days (SD 3.9) estimated from Li *et al.* [8].

Hence, based on the empirical distribution of delay from onset to report, we can construct the distribution for the delay from infection to report by convolution of incubation period distribution and the empirical distribution of delay from onset to report.

The infectiousness since infection $w_t$ was a convolution of incubation period and the infectiousness relative to onset (allowed to be pre-symptomatic) based on viral shedding data, with a shifted gamma distribution [9].

To account for the fact that imported cases did not contribute their entire infectiousness in Hong Kong, we assumed they started to contribute their infectiousness at onset in Hong Kong (the median of the difference between arrival date and onset date is 0 based on the "eCOVID-19" data). Hence, the infectiousness profile of local cases, $w_L(t) = w(t)$. For the infectiousness profile of imported cases, we assumed that it started at 5 days after infection since the mean incubation period is 5 days. Therefore, the infectiousness profile of imported cases is:

$$w_I(t) = I(t \geq 5) * \frac{w(t)}{\sum I(t \geq 5) w(t)}$$

We analyse the epidemic curve up to Apr 24, 2020, and take $\tau = 14$ in our analysis, to avoid unstable estimates for time-varying reproductive number.

In the dataset, local cases were further classified as unlinked local cases or local cases linked with other local cases. In our analysis, we sampled the source of unlinked local cases with equal probability to be infected from imported cases or local cases.

### 2.3 Inference

To inference $R_t$, we first use the deconvolution summarized in Section 2.2. An expectation–maximization (EM) algorithm with a smoothing step is also included to obtain a smoother epidemic curve to enhance interpretation. This is achieved by using the backprojNP function in the surveillance package in R.

After obtaining the epidemic curve by infection time, we use the model in Section 1.2. to estimate $R_t$ for local cases and imported cases. We use a Markov chain Monte Carlo approach to estimate the model parameter, as stated in Section 1.5.

We accounted for the uncertainty of input parameters, including incubation period and infectiousness profile to obtain the final estimates of $R_t$ in addition to model parameter uncertainty as follows:

We followed the approach in Salje et al. [10]. We reconstructed 200 epidemic curves. For each epidemic curve, we first sampled the parameters for the incubation period distribution. Then we used the deconvolution with that incubation period distribution and estimate $R_t$ for local cases and imported cases. Then we sampled the source of unlinked local cases. Then we presented the mean, 2.5% and 97.5% quantiles for those 200 $R_t$ estimates for each time point.

### 3 Reference

1. Fraser C. Estimating individual and household reproduction numbers in an emerging epidemic. PLoS One. 2007;2(8):e758. doi: 10.1371/journal.pone.0000758. PubMed PMID: 17712406; PubMed Central PMCID: PMCPMC1950082.
2. Cori A, Ferguson NM, Fraser C, Cauchemez S. A new framework and software to estimate time-varying reproduction numbers during epidemics. Am J Epidemiol. 2013;178(9):1505-12. doi: 10.1093/aje/kwt133. PubMed PMID: 24043437; PubMed Central PMCID: PMCPMC3816335.
3. Thompson RN, Stockwin JE, van Gaalen RD, Polonsky JA, Kamvar ZN, Demarsh PA, et al. Improved inference of time-varying reproduction numbers during infectious disease outbreaks. Epidemics. 2019;29:100356. Epub 2019/10/19. doi: 10.1016/j.epidem.2019.100356. PubMed PMID: 31624039; PubMed Central PMCID: PMCPMC7105007.
4. Gilks WR, Richardson S, Spiegelhalter D. Markov Chain Monte Carlo in Practice. London: Chapman & Hall; 1996.
5. Abbott S HJ, Thompson RN et al. Estimating the time-varying reproduction number of SARS-CoV-2 using national and subnational case counts [version 2; peer review: awaiting peer review]. Wellcome Open Res 2020, 5:112 (https://doi.org/10.12688/wellcomeopenres.16006.2).

6. Gostic KM, McGough L, Baskerville EB, Abbott S, Joshi K, Tedijanto C, et al. Practical considerations for measuring the effective reproductive number, Rt. PLoS Comput Biol. 2020;16(12):e1008409. Epub 2020/12/11. doi: 10.1371/journal.pcbi.1008409. PubMed PMID: 33301457; PubMed Central PMCID: PMCPMC7728287.

7. Becker NG, Watson LF, Carlin JB. A method of non-parametric back-projection and its application to AIDS data. Stat Med. 1991;10(10):1527-42. Epub 1991/10/01. doi: 10.1002/sim.4780101005. PubMed PMID: 1947509.

8. Li Q, Guan X, Wu P, Wang X, Zhou L, Tong Y, et al. Early Transmission Dynamics in Wuhan, China, of Novel Coronavirus-Infected Pneumonia. N Engl J Med. 2020. doi: 10.1056/NEJMoa2001316. PubMed PMID: 31995857.

9. He X, Lau EHY, Wu P, Deng X, Wang J, Hao X, et al. Temporal dynamics in viral shedding and transmissibility of COVID-19. Nat Med. 2020;26(5):672-5. doi: 10.1038/s41591-020-0869-5. PubMed PMID: 32296168.

10. Salje H, Cummings DAT, Rodriguez-Barraquer I, Katzelnick LC, Lessler J, Klungthong C, et al. Reconstruction of antibody dynamics and infection histories to evaluate dengue risk. Nature. 2018;557(7707):719-23. doi: 10.1038/s41586-018-0157-4. PubMed PMID: 29795354; PubMed Central PMCID: PMCPMC6064976.

### SUPPLEMENTARY FIGURE LEGEND

**Figure S1.** Simulation results for scenario 1: $R_t$ for imported cases (Panel A) and for local cases (Panel B) were set to be 1 and 1.5 respectively. The red lines indicated the true value of $R_t$, and the black lines indicated the estimates of $R_t$ and the blue lines indicated the 95% credible intervals for estimates.

**Figure S2.** Simulation results for scenario 2: $R_t$ for imported cases (Panel A) were set to be 1. $R_t$ for local cases (Panel B) were set to be 2.0 for the first 30 days and decreased to 0.8 for next 40 days. The red lines indicated the true value of $R_t$, and the black lines indicated the estimates of $R_t$ and the blue lines indicated the 95% credible intervals for estimates.
